# Functional variation in human Carbohydrate-Active enZYmes (hCAZymes) in relation to the efficacy of a FODMAP-reducing diet in IBS patients

**DOI:** 10.1101/2024.02.05.24302204

**Authors:** Andreea Zamfir-Taranu, Britt-Sabina Löscher, Florencia Carbone, Abdullah Hoter, Cristina Esteban Blanco, Isotta Bozzarelli, Leire Torices, Karen Routhiaux, Karen Van den Houte, Ferdinando Bonfiglio, Gabriele Mayr, Maura Corsetti, Hassan Y Naim, Andre Franke, Jan Tack, Mauro D’Amato

**Author notes:** **Guarantor of the article:** Mauro D’Amato, PhD, Department of Medicine and Surgery, LUM University, Casamassima, Italy. **Potential competing interests:** MD’A received consulting fees and MD’A and HYN received unrestricted research grants from QOL Medical LLC. The sponsor had no role in the study design or in the collection, analysis, and interpretation of data. **Specific author contributions:** MD’A and JT study design and supervision; B-SL, FC, KVH, AH, CEB, IB, LT, KR, GM, HYN, AF and JT data acquisition, patients characterization; AZT, B-SL and FB statistical and computational analyses; AZT, B-SL, FC, MC, HYN, AF, JT and MD’A data analysis and interpretation; HYN, AF, JT and MD’A obtained funding and technical support; AZT and MD’A drafted the manuscript, with input and critical revision from all other authors. All authors approved the final draft of the manuscript. **Patient consent for publication:** Not applicable. **Twitter** Andreea Zamfir-Taranu @ZamfirTaranu and Mauro D’Amato @damato_mauro.

## Abstract

**Objective:** Limiting the dietary intake of carbohydrates poorly absorbed in the small intestine (FODMAPs) has therapeutic effects in some but not all irritable bowel syndrome (IBS) patients. We investigated genetic variation in human Carbohydrate-Active enZYmes (hCAZymes) genes in relation to the response to a FODMAP-lowering diet in the DOMINO study.

**Design:** hCAZy polymorphism was studied in IBS patients from the dietary (FODMAP-lowering; N=196) and medication (otilonium bromide; N=54) arms of the DOMINO trial via targeted sequencing of 6 genes of interest (*AMY2B*, *LCT*, *MGAM*, *MGAM2*, *SI* and *TREH*). hCAZyme defective (hypomorphic) variants were identified via computational annotation using clinical pathogenicity classifiers. Age-and sex-adjusted logistic regression was used to test hCAZyme polymorphisms in cumulative analyses where IBS patients were stratified into carrier and non-carrier groups (collapsing all hCAZyme hypomorphic variants into a single bin). Quantitative analysis of hCAZyme variation was also performed, in which the number of hCAZyme genes affected by a hypomorphic variant was taken into account.

**Results:** In the dietary arm, the number of hypomorphic hCAZyme genes positively correlated with treatment response rate (P=0.03, OR=1.51 [CI=0.99-2.32]). In the IBS-D group (N=55), hCAZyme carriers were six times more likely to respond to the diet than non-carriers (P=0.002, OR=6.33 [CI=1.83-24.77]). These trends were not observed in the medication arm.

**Conclusions:** hCAZYme genetic variation may be relevant to the efficacy of a carbohydrate-lowering diet. This warrants additional testing and replication of findings, including mechanistic investigations of this phenomenon.

**WHAT IS KNOWN:**

- Carbohydrates are known triggers of IBS symptoms, and limiting their dietary consumption appears to have therapeutic effects
- The lowFODMAP diet improves symptoms in some but not all IBS patients

**WHAT IS NEW HERE:**

- Carrying hypomorphic hCAZyme gene variants associates with increased efficacy of a FODMAP-lowering diet in IBS, especially in patients with diarrhoea (IBS-D)
- hCAZyme genotype information may be relevant to increase therapeutic precision in IBS, contributing to personalising carbohydrate-focused dietary interventions

## INTRODUCTION

Irritable Bowel Syndrome (IBS) is a disorder of gut-brain interaction (DGBI) affecting 5-10% of the general population with recurrent symptoms including abdominal pain, bloating, diarrhea (IBS-D), constipation (IBS-C), or a combination of diarrhea and constipation (IBS-M).[1-4]The pathophysiology of IBS is multifactorial, contributed by several risk factors: psychological stressors, prior infections, gut dysbiosis, epithelial barrier dysfunction, mucosal immunity and dietary triggers.[5,6] The latter in particular might explain some of the clinical manifestations associated with postprandial IBS symptoms,[7] and avoidance of carbohydrates (owing to their potential maldigestion) is recommended as a first-line approach by the UK England and Wales National Institute for Health and Care Excellence (NICE) guidelines for IBS. Limiting the dietary consumption of certain carbohydrates poorly absorbed in the small intestine (known as fermentable oligosaccharides, disaccharides, monosaccharides and polyols-FODMAPs) also appears to have therapeutic effects in some but not all IBS patients.[8-14] The food–symptom relation in IBS may involve maldigestion of carbohydrates due to inefficient enzymatic breakdown of polysaccharides, which can be fermented by colonic microbiota generating gas and short-chain fatty acids ultimately affecting gastrointestinal function.[15-17]

Polysaccharide breakdown is a process that, in humans, involves the action of several Carbohydrate-Active enZYmes (hCAZymes, http://www.cazy.org/e355.html) from the upper GI tract and the small intestine (Figure S1). Carbohydrate digestion is initiated by salivary and pancreatic amylases (AMYs) and then finalised in the small intestine by other glycosidases (brush-border enzymes like lactase - LCT, sucrase-isomaltase - SI, maltase-glucoamylase - MGAM and trehalase - TREH) that hydrolyze disaccharides into monosaccharides ultimately absorbed by the enterocytes. Highlighting this, hCAZyme mutations cause rare genetic forms of carbohydrate intolerance, while regulatory DNA variations (persistence genotype) influence lactose intolerance in adults.[18,19]

While genetic forms of carbohydrate maldigestion present with clinical manifestations overlapping with (and sometimes misdiagnosed as) IBS,[20] compelling evidence for similar mechanisms underlying bowel symptoms in a subset of IBS patients is accumulating, mostly in relation to the *SI* gene: in a series of case-control and population-based studies *SI* hypomorphic variants with (experimentally verified and/or computationally predicted) reduced enzymatic activity have been shown to confer an increased risk of IBS and,[17,21-24] respectively, decreased and increased chances to respond to low-FODMAP and sucrose-and starch-reduced (SSRD) diets.[25,26] This suggests similar mechanisms may be detected for other hCAZymes involved in the breakdown of FODMAPs and other carbohydrates found in foods that are typically restricted in carbohydrates-focused diets. We sought to test this hypothesis in a pilot analysis of the correlation between hCAZyme gene carrier status and response to a FODMAPs-lowering intervention in the Diet Or Medication in Irritable Bowel syNdrOme (DOMINO) trial. The outcome of this type of studies may have implications in the management of IBS, by means of allowing the design of new approaches for personalizing dietary intervention in a subset of genetically exposed IBS patients.

## MATERIALS AND METHODS

### Patients cohort

The current study is based on retrospective (genetic) analyses conducted on IBS patients from the DOMINO trial (NCT04270487), previously described. [27] Briefly, Belgian IBS patients (N=459) diagnosed by primary care physicians (including IBS-M, IBS-D, and IBS-C subtyping based on stool type) were randomised to one of two 8 week treatments: a diet app designed to lower the intake of FODMAPs (dietary arm; N=218) or 40 mg 3x/day of the antispasmodic agent otilonium bromide (OB; medication arm; N=217). [27] Based on data from IBS Symptom Severity Score (IBS-SSS) questionnaires, a positive response was defined as a drop of 50 IBS-SSS points, compared to baseline. For the purpose of hCAZyme genetic analyses, and after quality control of Illumina targeted next-generation sequencing (NGS) and Global Screening Array (GSA) genotyping data (see below), 196 IBS patients from the dietary arm (39 IBS-C, 55 IBS-D, 73 IBS-M and 29 IBS-U), were deemed eligible for analyses and were included in this study. Additionally, 54 IBS-D patients from the medication arm were also included in parallel (negative control) analyses. The demographics, clinical characteristics, stool type, IBS-SSS profiles, and response to treatment of these patients are reported in Table 1 (additional data, including adherence and satisfaction with the diet, quality of life, and extra-intestinal symptoms are reported in the original publication).[27]

**Table 1.** Demographic and clinical characteristics of patients included in the study.

|  | **Diet\*** | **Medication\*** |
| --- | --- | --- |
| Patients: N | 196 | 54 |
| Females: N (%) | 147 (75.0) | 38 (70.4) |
| Age: mean ± SD | 41.0 ± 14.7 | 37.5 ± 14.3 |
| Response: N (%) | 138 (70.4) | 34 (63.0) |
| **Stool type** |  |  |
| IBS-D: N (%) | 55 (28.1) | 54 (35.3) |
| IBS-M: N (%) | 73 (37.2) | - |
| IBS-C: N (%) | 39 (19.9) | - |
| IBS-U: N (%) | 29 (14.8) | - |
| **Symptoms at baseline** |  |  |
| IBS-SSS score: mean ± SD | 270.4 ± 94.1 | 262.0 ± 97.0 |
| Abdominal distention severity: mean ± SD | 53.4 ± 25.9 | 46.7 ± 28.1 |
| Life disruption: mean ± SD | 50.6 ± 26.1 | 52.8 ± 25.2 |
| Abdominal pain severity: mean ± SD | 49.6 ± 24.7 | 51.5 ± 24.4 |
| Bowel habit satisfaction: mean ± SD | 67.5 ± 23.4 | 66.5 ± 26.0 |
| Abdominal pain duration (x10 days): mean ± SD | 4.9 ± 3.0 | 4.5 ± 2.9 |
| **Symptoms after 8 weeks treatment** |  |  |
| IBS-SSS score: mean ± SD | 172.5 ± 111.2 | 173.1 ± 99.7 |
| Abdominal distention severity: mean ± SD | 34.1 ± 27.3 | 33.9 ± 27.0 |
| Life disruption: mean ± SD | 29.3 ± 26.9 | 31.0 ± 23.4 |
| Abdominal pain severity: mean ± SD | 34.8 ± 24.7 | 37.2 ± 23.2 |
| Bowel habit satisfaction: mean ± SD | 43.9 ± 27.0 | 43.7 ± 23.8 |
| Abdominal pain duration (x10 days): mean ± SD | 3.1 ± 2.6 | 3.3 ± 2.3 |

\* No significant difference detected for any of the variables reported.

### Ethics Statement

The study protocol was approved by the Ethical Committee Research UZ/KU Leuven (protocol S59482), and all participants provided informed consent.

### Targeted sequencing of hCAZyme genes

An Illumina AmpliSeq DNA targeted next-generation sequencing (NGS) assay focused on IBS-candidate genes was designed using Illumina Design Studio, which included ten hCAZyme genes (*AMY1A*, *AMY1B*, *AMY1C*, *AMY2A*, *AMY2B*, *LCT*, *MGAM*, *MGAM2*, *SI*, *TREH*).[28] Panel sequencing was carried out on DNA from selected DOMINO participants and, for the purpose of this study, high-quality sequencing data were further analysed for six genes (*AMY2B*, *LCT*, *MGAM*, *MGAM2*, *SI* and *TREH)* whose entire coding sequence (all exons and exon boundaries) was successfully covered (>90% of the target sequence with 30x coverage). Sequencing of 2×150 bp paired-end reads was performed on an Illumina NextSeq, and the reads mapped to the reference human genome build GRCh38 using the IKMB exome pipeline (https://github.com/ikmb/exome-seq). Variant calling was performed using DeepVariant v.1.0. [29] and Genome Analysis Toolkit (GATK),[30] in parallel and only variants called by both tools were included in the analyses.[31] Targeted sequencing data were further validated by whole exome sequencing on 5 individuals randomly selected among DOMINO participants, obtaining identical results. ANNOVAR [32] was used for functional annotation of hCAZyme variants, in order to filter for coding missense (non-synonymous and/or stop-gain/loss) changes, before assigning pathogenicity scores (see below). GSA genotyping data available for DOMINO participants [33] was used for cohort-level quality controls, filtering our individuals of non-European ancestry, with missing data, heterozygosity rate >6SD and/or calling rate <0.85.

### Identification of hCAZyme hypomorphic variants

To identify hCAZyme hypomorphic variants (defective variants with absent or reduced enzymatic activity), the functional relevance of non-synonymous changes was predicted *in silico* using pathogenicity scores derived from a combination of the M-CAP [34] and CADD [35] computational tools, as previously described.[22]

### Statistical Analyses

Statistical analyses were performed with R v.4.2.1 [36] in RStudio (v2022.07.2+576). R packages ggplot2 [37] and dplyr [38] were used for data visualisation and manipulation. hCAZyme hypomorphic variants were tested versus the response to treatment in cumulative analyses, collapsed into a single hypomorphic hCAZyme group. Accordingly, DOMINO participants were defined as carriers or non-carriers of any hCAZyme gene hypomorphic variant, while the number of hCAZyme genes affected by hypomorphic variants was also taken into account in separate analyses (Figure 1). hCAZyme carrier status (independent variable, binary) and number of hCAZymes affected genes (independent variable, continuous) were tested for their effect on treatment response (dependent variable, binary) using logistic regression (one-tail) adjusting for participants’ sex and age. Similar analyses were carried out for IBS subtypes. Fisheŕs Exact test was used to test *SI* and hCAZyme variants in sensitivity analyses due to small sample sizes and the presence of 0 values in the outcome variable groups.

**Figure 1.**
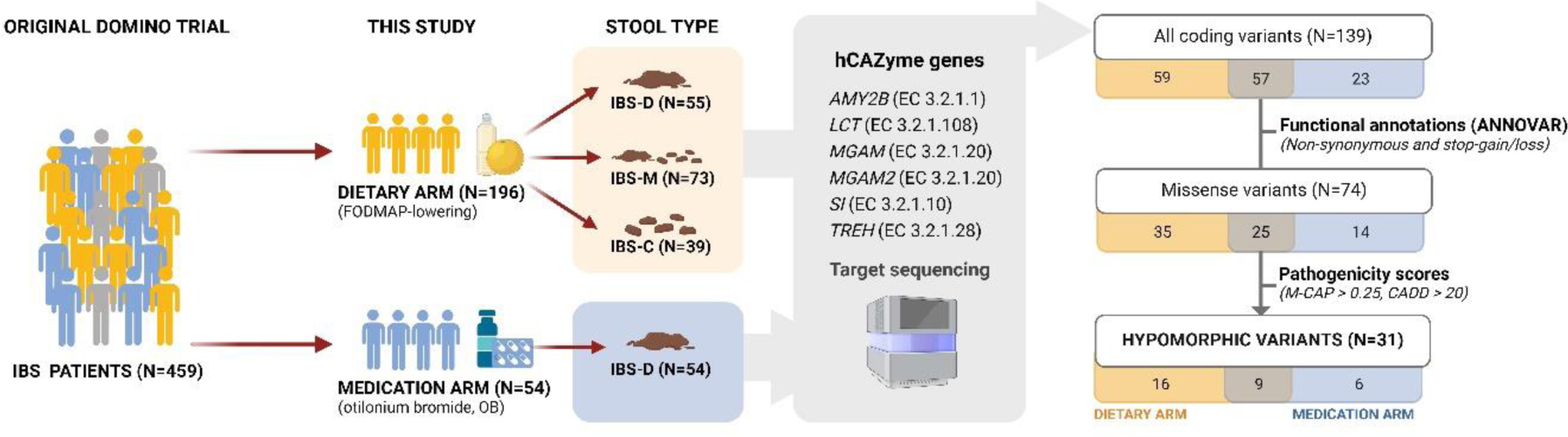
Graphical representation of the methodology used in this study (flowchart).

## RESULTS

### Selection of hCAZyme genes and genomic sequencing

The annotation of the molecular pathways *Carbohydrate digestion and absorption* (https://www.kegg.jp/entry/map04973) and *Starch and sucrose metabolism* (https://www.kegg.jp/entry/map00500) from the Kyoto Encyclopedia of Genes and Genomes (KEGG) was used to select hCAZyme genes to be included in an IBS-focused targeted NGS panel (see Methods). Upon implementation of this assay to sequence DOMINO participants’ DNA (see below), high-coverage/high-quality NGS data could be obtained for the hCAZyme genes *AMY2B*, *LCT*, *MGAM, MGAM2, SI* and *TREH*, which were included in downstream analyses.

### Identification of hCAZyme hypomorphic variants

After sample and NGS quality controls (see Methods), a total of 196 DOMINO patients from the dietary arm were deemed eligible for inclusion in the genetic analyses. In these individuals, we detected a total of 60 missense hCAZyme variants (see Methods), all previously identified in other populations (present in the gnomAD genome reference database) (Figure 1).[39] These included 49 rare and 11 common hCAZyme variants (allele frequencies respectively below or above 5% in gnomAD), which are reported in Table S1. Based on computational predictions of the functional effects of these amino acid changes (pathogenicity scores from M-CAP and CADD, see Methods), 25 hCAZyme variants were classified as hypomorphic (dysfunctional), the remaining 35 as benign. Most hypomorphic variants were rare and only found in single individuals, while two were common and detected in 18 (Leu1174Met rs73547325 SNP in *MGAM2*) and 113 (Val15Phe, rs9290264 SNP in *SI*) patients, respectively (Table 2, Figure S2).

**Table 2.** hCAZyme hypomorphics variants identified in this study.

| Gene | Variant | dbSNP | Ref allele | Alt allele | gnomAD freq | freq this study | Carriers Diet (N=196) | Carriers OB (N=54) |
| --- | --- | --- | --- | --- | --- | --- | --- | --- |
| **AMY2B** |  |  |  |  |  |  |  |  |
|  | Val190Leu | - | G | T | 0.0000009 | 0.002 | - | 1 |
|  | Val309Ala | rs140209167 | T | C | 0.0036 | 0.006 | 2 | 1 |
|  | Gly319Arg | rs140978983 | G | A | 0.0315 | 0.044 | 16 | 6 |
|  | Ile406Thr | rs151132065 | T | C | 0.0220 | 0.018 | 7 | 2 |
| **LCT** |  |  |  |  |  |  |  |  |
|  | Gly928Cys | - | C | A | - | 0.002 | 1 | - |
|  | Asn1035Tyr | - | T | A | - | 0.004 | 2 | - |
| **MGAM** |  |  |  |  |  |  |  |  |
|  | Arg401Cys | rs188481752 | C | T | 0.0032 | 0.002 | 1 | - |
|  | Arg637Ser | rs190777514 | A | C | 0.0019 | 0.002 | 1 | - |
|  | Leu806Ile | rs956495934 | C | A | 0.00002 | 0.002 | 1 | - |
|  | Leu854Phe | rs200141280 | C | T | 0.0013 | 0.002 | - | 1 |
|  | Asn858Asp | rs2960746 | A | G | 0.0001 | 0.006 | 3 | - |
|  | Cys873Trp | rs768578658 | T | G | 0.0001 | 0.002 | 1 | - |
|  | Pro1424Thr | rs185053832 | C | A | 0.0108 | 0.010 | 3 | 2 |
|  | Leu1794Ser | rs201177568 | T | C | 0.0010 | 0.002 | 1 | - |
| **MGAM2** |  |  |  |  |  |  |  |  |
|  | Ile113Thr | - | T | C | 0.000006 | 0.002 | - | 1 |
|  | Asn606Ser | rs79591013 | A | G | 0.0122 | 0.012 | 4 | 2 |
|  | Cys619Arg | rs774919724 | T | C | 0.0006 | 0.002 | - | 1 |
|  | Ala1138Val | - | C | T | - | 0.002 | 1 | - |
|  | Leu1174Met | rs73547325 | T | A | 0.0749 | 0.052 | 18 | 8 |
|  | Gln1546Glu | rs7776662 | C | G | 0.00002 | 0.002 | - | 1 |
|  | Val2018Ile | - | G | A | - | 0.016 | 8 | - |
| **SI** |  |  |  |  |  |  |  |  |
|  | Met1Ile | - | C | T | - | 0.002 | 1 | - |
|  | Val15Phe | rs9290264 | C | A | 0.2990 | 0.340 | 113 | 30 |
|  | Val371Met | rs138434001 | C | T | 0.0032 | 0.004 | 2 | - |
|  | Arg774Gly | rs147207752 | T | C | 0.0015 | 0.004 | 2 | - |
|  | Ile799Val | rs150246328 | T | C | 0.0042 | 0.004 | 1 | 1 |
|  | Tyr975His | rs146785675 | A | G | 0.0055 | 0.002 | 1 | - |
|  | Ser1490Ile | rs376437234 | C | A | 0.0001 | 0.002 | - | 1 |
|  | Leu1520Ter | - | - | A | - | 0.008 | 1 | 3 |
| **TREH** |  |  |  |  |  |  |  |  |
|  | Lys140Arg | rs34978247 | T | C | 0.0069 | 0.006 | 3 | - |
|  | Met512Thr | rs556006762 | A | G | 0.0003 | 0.002 | 1 | - |

**Gene:** gene name; **Variant:** corresponding amino acid change; **dbSNP:** dbSNP database ID; **Ref Allele:** reference allele according to gnomAD v3.1.1 data (https://gnomad.broadinstitute.org/); **Alt Allele:** alternative allele according to gnomAD; **gnomAD freq:** allele frequency in individuals of non-Finnish European ancestry, as from gnomAD; **freq this study:** allele frequency in 250 IBS DOMINO samples from dietary and medication arm; **Carriers Diet:** number of carriers among 196 IBS patients from the dietary arm; **Carriers OB:** number of carriers among 54 IBS-D patients from the OB arm. "-": data not available.

For statistical analyses, hCAZyme hypomorphic variants were collapsed into a single group, and individuals were classified as carriers or non-carriers depending on whether they had at least one hypomorphic variant in one or more of the hCAZyme genes (carriers), or no hypomorphic variant (non-carriers). In addition, hCAZyme carrier status was evaluated under a gene-dosage model in relation to the number of hCAZyme genes affected by the presence of hypomorphic variants (irrespective of the number of variants present in each gene).

### hCAZyme genes and response to dietary treatment

DOMINO IBS patients treated with the FODMAP-lowering diet were stratified in hCAZyme carrier and non-carrier groups, and their corresponding clinical characteristics are reported in Table S2. While their demographic and clinical characteristics were similar, hCAZyme carriers generally exhibited more severe symptoms at baseline (Figure S3), especially in the IBS-D group where significant associations were detected both for total IBS-SSS and individual symptoms scores (higher in single and multiple hCAZy carriers compared to non-carriers; Figure 2). When the outcome of the FODMAP-lowering diet was studied in a sex-and age-adjusted logistic regression (see Methods), a significant association between hypomorphic hCAZyme gene number and response rate was detected: the diet was more effective in patients with multiple (computationally predicted) defective hCAZyme genes (P=0.03, OR=1.51 [CI 0.99-2.32]), reaching a 100% success rate in the small group of IBS patients carrying defective variants in three hCAZyme genes (Figure 3). A similar pattern was also observed at the level of severity of individual symptoms (from IBS-SSS), which tended to correlate with the number of affected hCAZyme genes (Figure S4). Near-significant findings were also obtained for carriers of hypomorphic hCAZyme genes who were more likely to respond to the dietary treatment (P=0.06, OR=1.69 [CI 0.87-3.29]; Figure S5). As shown in Figure 4, this phenomenon was most pronounced in the IBS-D subtype, with hCAZyme carriers >6x more likely to respond to the FODMAP-reducing diet than non-carriers as assessed on the IBS-SSS severity scale both at the global (P=0.002, OR=6.33 [CI=1.83-24.77]) and, as a tendency, individual symptom level (Figure S6). No significant findings were detected for the IBS-M and IBS-C subtypes (not shown).

**Figure 2.**
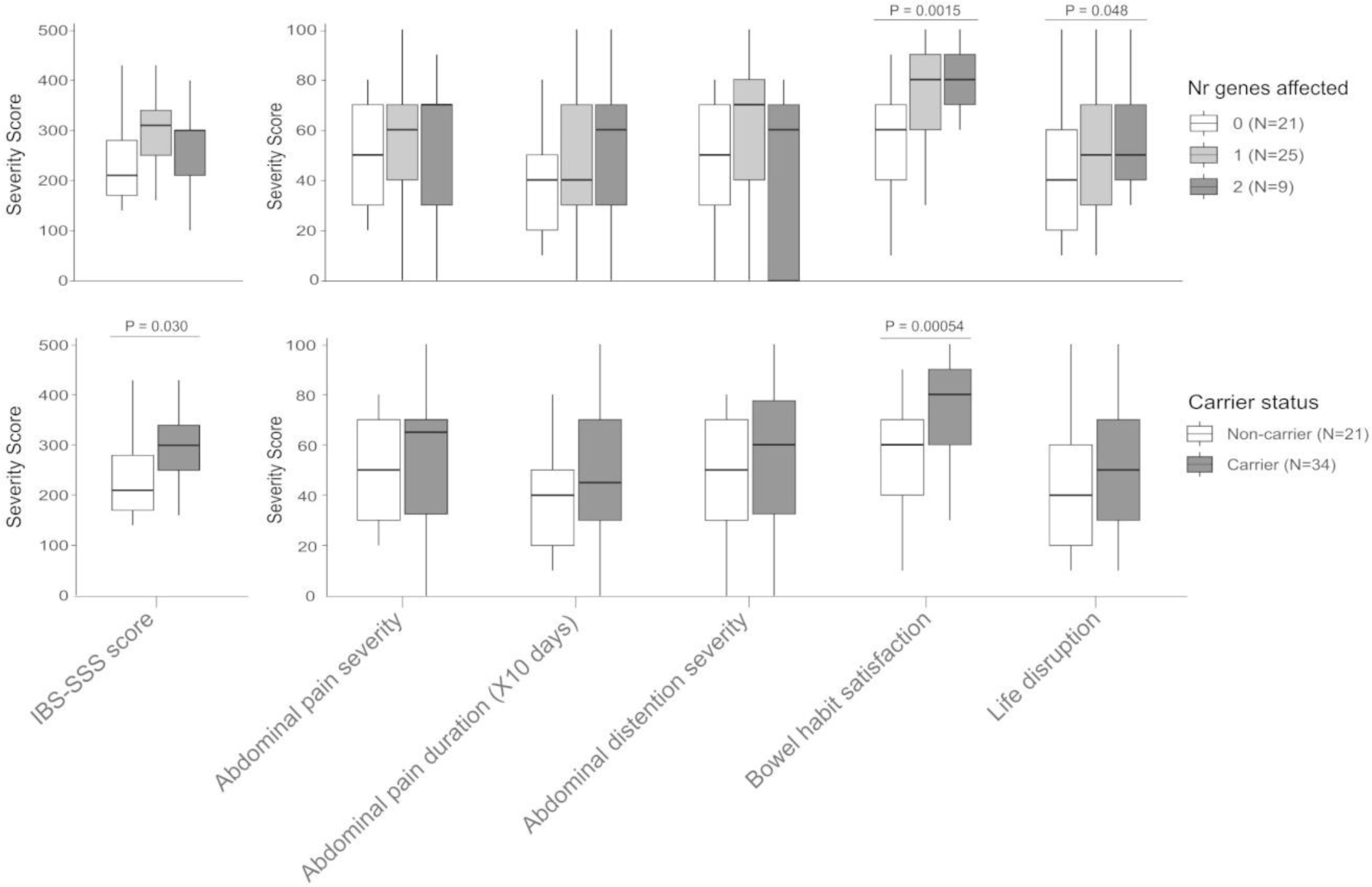
Baseline IBS symptoms scores in for 55 IBS-D DOMINO patients from the dietary arm stratified according to genetic variation in the hCAZyme genes: top) number of affected hCAZyme genes; bottom) hCAZyme carrier status. p=0.048, p=0.030, p=0.0015 and p=0.00054 (linear regression adjusted for age+sex, one-tail).

**Figure 3.**
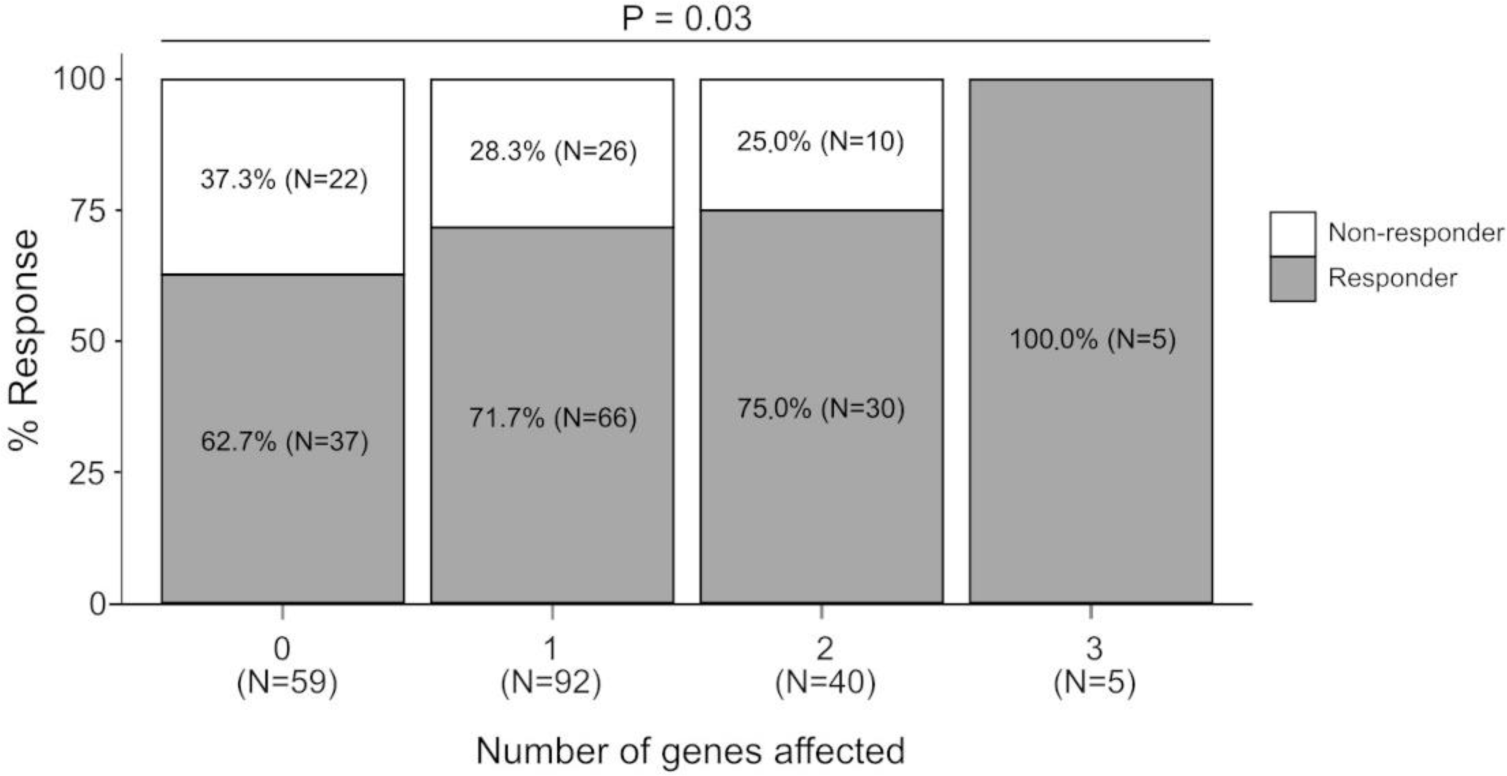
Response to FODMAP-lowering diet in 196 IBS DOMINO patients, stratified according to the number of hCAZyme genes affected. p=0.03 (logistic regression adjusted for age+sex, one-tail).

**Figure 4.**
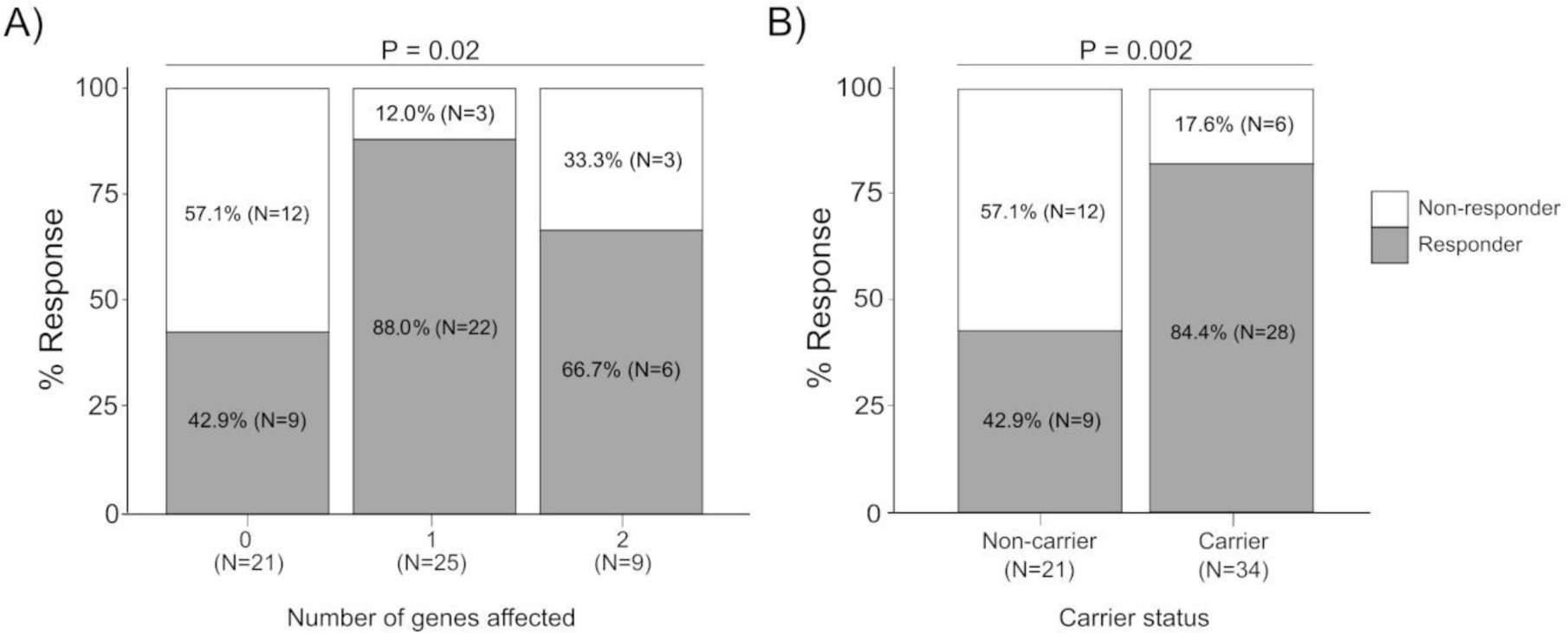
Response to FODAMP-lowering diet in 55 IBS-D DOMINO patients, stratified according to A) the number of hCAZyme genes affected and B) the hCAZyme carrier status. p=0.02 and p=0.002 (logistic regression adjusted for age+sex, one-tail).

### *SI* sensitivity analyses

*SI* hypomorphic variants (particularly the common Val15Phe variant) were the most common hCAZyme variants in DOMINO, and have been previously shown to affect the response to a low-FODMAP diet,[25] hence we ran additional sensitivity analyses. There was no significant association with the response to dietary treatment when the whole DOMINO cohort was stratified into *SI* hypomorphic carriers and non-carriers (irrespective of other hCAZyme variants, not shown), and excluding *SI* carriers from the analyses confirmed near-significant higher response rate in hCAZyme carriers despite sample size reduction (N=81; 18/22=81.8% and 37/59=62.7% response rate in hCAZyme carriers vs non-carriers, respectively; P=0.06). Finally, in the small group of IBS-D patients (N=27), *SI*-devoid hCAZyme hypomorphic variants were again significantly associated with increased response to the FODMAP-lowering diet (6/6=100% vs 9/21=42.9 response rate in hCAZyme carriers vs non-carriers, respectively, P=0.01). Overall, this indicates hCAZyme findings are not dependent (only) on *SI* genotype.

### hCAZyme genes and response to medication

To further explore the relationship between hypomorphic hCAZyme carrier status and response to treatment, we performed additional analyses in the group of patients who were administered OB, that is in a situation where alteration of carbohydrate digestion may not be relevant. Given previous findings in the IBS-D group of patients, where hCAZyme genotype appeared to be most relevant, we selected OB-treated IBS-D patients to run similar analyses. Genomic sequencing identified 15 hypomorphic hCAZyme variants in this group (Table 2). While no significant relationship was observed with the number of affected hCAZyme genes (Figure S7), OB treatment even proved to be significantly less effective in hypomorphic hCAZyme carriers than non-carriers (Figure 5), thus showing a pattern opposite to the dietary arm of the DOMINO trial.

**Figure 5.**
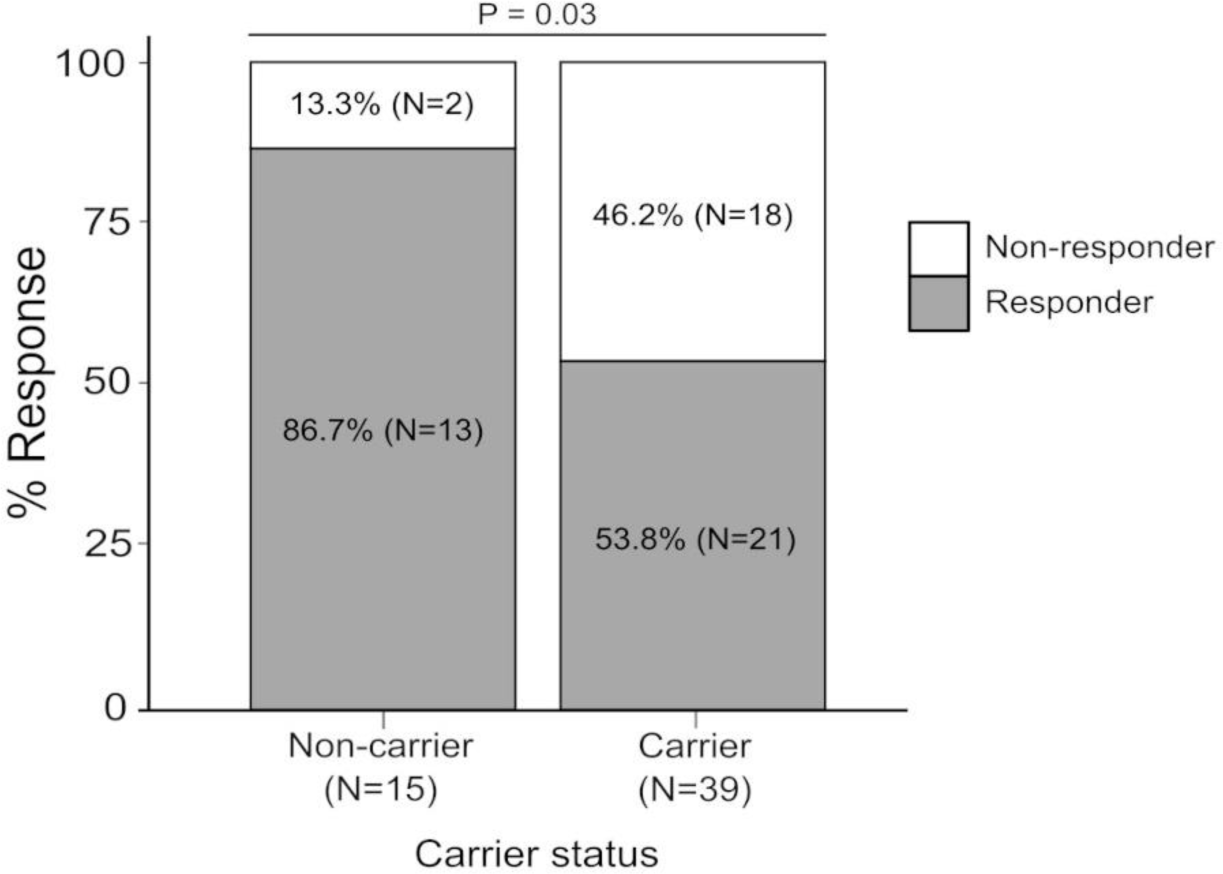
Response to OB treatment in 54 IBS-D DOMINO patients, stratified according to hCAZyme carrier status. p=0.03 (logistic regression adjusted for age+sex, one-tail).

## DISCUSSION

We report here a first survey of IBS treatment response in relation to genetic variation in genes involved in the breakdown and digestion of dietary carbohydrates. We show that the hypomorphic hCAZyme genotype is associated with an increased likelihood of responding to a FODMAP-reducing diet while being irrelevant (if not deleterious) against a treatment based on medication that affects unrelated, different biological mechanisms (OB is an L-type calcium blocker acting as a spasmolytic agent on intestinal smooth muscles).

In a retrospective analysis of the two-arm dietary and medication DOMINO interventional trial, we focused our genetic analyses on glycoside hydrolases (EC 3.2.1 enzymes AMY2B, LCT, MGAM, MGAM2, SI and TREH), the battery of hCAZy enzymes responsible for the digestion of most dietary sugars and starch. Recessive mutations in the corresponding genes are known to cause bowel symptoms in several forms of carbohydrate intolerance (lactose, sucrose, trehalose), due to the migration of undigested saccharides to the lower bowel where they are fermented by resident bacteria inducing gas production, bloating and diarrhoea.[15,24,40] This represents the foundation of the hypothesis tested in our study: individuals with dysfunctional (hypomorphic) hCAZyme alleles may benefit more than wild-type carriers from diets restricting carbohydrate intake (like it ensues in a FODMAP-reducing diet), possibly in a gene-dosage fashion in which multiple-hCAZyme carriers are best responders. Focusing on IBS also finds rationale in previous surveys showing how hypomorphic *SI* variants can affect both the risk of IBS and the response to low-FODMAP and SSRD carbohydrate-reducing diets.[16,17,22-26]

The FODMAP-lowering diet implemented in the DOMINO trial limits the consumption of a series of carbohydrates (FODMAPs and others contained in the restricted foods) that are substrates for hCAZy enzymes predictably defective in hypomorphic variant carriers, and these patients (especially those with multiple hCAZymes affected) appear to benefit more from this dietary restriction than non-carriers. Of note, this evidence was most pronounced in the IBS subtype characterized by diarrhoea (IBS-D), which is a clinical manifestation typical of carbohydrate maldigestion. Taken together, these data also suggest that, similar to rare forms of carbohydrate maldigestion, functional changes in CAZyme genes might contribute to generate bowel symptoms via carbohydrate maldigestion in IBS.[20]

If confirmed in follow-up studies, the results reported here may have implications in the management of IBS (especially IBS-D) because of the potential for improving the specificity and efficacy of dietary intervention, providing a rationale for personalised therapeutic (dietary) approaches based on hCAZymes genotype. Indirectly, this may also have positive repercussions on patients’ adherence and compliance. As recent surveys show, adherence to a low-FODMAP diet is difficult and requires considerable effort from the patient including several visits to a dietitian, with a significant proportion of IBS patients failing to respond because of non-compliance.[41,42] Prior knowledge of hCAZyme genotype (and associated likelihood of benefiting from the diet) may thus contribute to decision-making and the ultimate propensity of a patient to agree to adopt this or similar dietary intervention schemes.

The exact mechanism(s) by which hCAZyme genotype influences the likelihood to respond to a carbohydrate-(FODMAP-) reducing diet was not experimentally verified, as this was beyond the scope of this observational study. However, it is reasonable to assume it is rooted in the accumulation of undigested (poly-, oligo-and di-) saccharides in the lower bowel of hCAZyme hypomorphic carriers, where they are fermented by the resident bacteria. Indeed, dietary carbohydrates are known to have an important role in shaping the gut microbiota, with bacteria carrying prokaryotic CAZyme genes showing a competitive advantage over others when undigested polysaccharides are abundant in the large bowel.[43,44] This is relevant to IBS, where the microbiota plays an important role, and where symptoms and their severity have been shown to correlate with bacterial carbohydrate metabolism among other factors.[45-48] Future studies are warranted where the relationship between hCAZyme genotype and gut microbiota composition is investigated.

Additional studies are also needed to elucidate the relationship between individual hCAZyme genes and specific saccharides ultimately impacting the response to dietary treatment, since the relatively small DOMINO sample size and the fact that several individuals were positive at multiple hCAZyme loci did not allow meaningful analyses to be carried out at the single hCAZyme gene level. The diet adopted in the DOMINO study was not a strict low FODMAP diet, but rather a FODMAP-lowering diet complemented with recommendations from the NICE guidelines for IBS; it did not foresee the use of food diaries allowing to quantify intake of individual carbohydrates. It should now be possible to design follow-up studies in which dietary restrictions of specific carbohydrates can be tested for their effect on symptom amelioration in carriers of individual hCAZyme genes directly involved in their digestion.

An important limitation of this study is that its findings could not be replicated in independent cohorts. While most CAZYme variants are rare and require suitable sample size to be tested, we are not aware of any other similarly-sized (N=196 from the dietary arm of the DOMINO study tested here) IBS dietary intervention trial that would allow meaningful validation efforts to be carried out. Future studies can be specifically planned for this purpose. At the same time, it is noteworthy that the analysis of hCAZyme genes in IBS-D patients from the medication arm did not reveal any association, which serves as an ideal negative control (that is, no association detected when the targeted mechanism is unrelated to CAZYme function).

Finally, the establishment of scalable, sensitive functional assays for hCAZyme relative gene variant activity quantification is needed, as past and current genetic research is mostly based on computational predictions that require validation in reproducible high-throughput *in vitro* systems.

In conclusion, we provide initial evidence that functional (hypomorphic) hCAZymes variations collectively influence the likelihood of responding to dietary treatment based on restricting the intake of certain carbohydrates. This information may be relevant to increase therapeutic precision and efficacy via genotype-driven dietary intervention in IBS. Future studies can be designed to consolidate this evidence with replication in independent cohorts and to obtain biological insight at the mechanistic level.

## Data Availability

All data produced in the present study are available upon reasonable request to the authors.

## ABBREVIATIONS

AMY: (amylase)
CADD: (Combined Annotation Dependent Depletion)
DGBI: (Disorder of Gut-Brain Interaction)
DOMINO: (Diet Or Medication in Irritable Bowel syndrome)
FODMAPs: (Fermentable Oligosaccharides, Disaccharides, Monosaccharides and Polyols)
GATK: (Genome Analysis Toolkit)
GSA: (Global Screening Array)
hCAZymes: (Human Carbohydrate-Active enZYmes)
IBS: (Irritable Bowel Syndrome)
IBS-SSS: (IBS-Symptom Severity Score)
KEGG: (Kyoto Encyclopedia of Genes and Genomes)
M-CAP: (Mendelian-Clinically Applicable Pathogenicity)
MGAM: (Maltase-GlucoAMylase)
NGS: (Next-Generation Sequencing)
OB: (Otilonium Bromide)
SI: (Sucrase-Isomaltase)
SNP: (Single Nucleotide Polymorphism)
SSRD: (Sucrose-and Starch-Reduced Diet)
TREH: (Trehalase)

## SUPPLEMENTARY MATERIAL FIGURE/TABLE LEGENDS

### FIGURES

**Figure S1.** Graphical representation of the process of carbohydrate digestion in the gastrointestinal system, including defects in hCAZYmes.

**Figure S2.** UpSet plot showing all genotypic combinations of hypomorphic variants present in hCAZyme genes from this study.

**Figure S3.** Baseline IBS symptoms in 196 IBS DOMINO patients from the dietary arm stratified according to genetic variation in the hCAZyme genes: top) number of affected hCAZyme genes; bottom) hCAZyme carrier status.

**Figure S4.** Variation of IBS symptoms after 8 weeks of treatment in 196 IBS patients from the dietary arm. Total (A) and individual (B-F) symptom scores are stratified according to genetic variation in the hCAZyme genes: left) number of affected hCAZyme genes; right) hCAZyme carrier status.

**Figure S5.** Response to a FODAMP-lowering diet in 196 IBS DOMINO patients stratified according to hCAZyme carrier status.

**Figure S6.** Variation of IBS symptoms after 8 weeks of treatment in 55 IBS-D patients from the dietary arm. Total (A) and individual (B-F) symptom scores are stratified according to genetic variation in the hCAZyme genes: left) number of affected hCAZyme genes; right) hCAZyme carrier status

**Figure S7.** Response to OB treatment in 54 IBS-D IBS DOMINO patients stratified according to the number of hCAZyme genes affected.

### TABLES

**Table S1.** hCAZyme variants identified in this study.

**Table S2.** Demographics and symptom scores in hCAZyme carrier groups from the dietary arm.

## Notes

**Financial support: Supported by funding from: the Spanish Government MCIN/AEI/10.13039/501100011033 (PCI2021-122064-2A to MD’A)**, the German Federal Ministry for Education and Research BMBF (01EA2208B to HYN and 01EA2208A to AF) and the Medical Research Council MRC (MR/W031213/1 to MC), under the umbrella of the European Joint Programming Initiative “A Healthy Diet for a Healthy Life” (JPI HDHL) and of the ERA-NET Cofund ERA-HDHL (GA N° 696295 of the EU Horizon 2020 Research and Innovation Programme); the Spanish Government MCIN/AEI/10.13039/501100011033 (PID2020-113625RB-I00 to MD’A); the German Research Foundation DFG (NA331/13-1 to HYN and 390884018 for the Excellence Cluster “Precision Medicine in Chronic Inflammation” to AF); the DOMINO study was supported by the Belgian Health Care Knowledge Centre (ref number: 16001).

### Competing Interest Statement

Mauro D Amato received consulting fees and Mauro D Amato and Hassan Y Naim received unrestricted research grants from QOL Medical LLC. The sponsor had no role in the study design or in the collection, analysis, and interpretation of data.

### Clinical Trial

NCT04270487

### Funding Statement

Supported by funding from: the Spanish Government MCIN/AEI/10.13039/501100011033 (PCI2021-122064-2A to MD A), the German Federal Ministry for Education and Research BMBF (01EA2208B to HYN and 01EA2208A to AF) and the Medical Research Council MRC (MR/W031213/1 to MC), under the umbrella of the European Joint Programming Initiative (A Healthy Diet for a Healthy Life) (JPI HDHL) and of the ERA-NET Cofund ERA-HDHL (GA No. 696295 of the EU Horizon 2020 Research and Innovation Programme); the Spanish Government MCIN/AEI/10.13039/501100011033 (PID2020-113625RB-I00 to MD A); the German Research Foundation DFG (NA331/13-1 to HYN and 390884018 for the Excellence Cluster (Precision Medicine in Chronic Inflammation) to AF); the DOMINO study was supported by the Belgian Health Care Knowledge Centre (ref number: 16001).

### Author Declarations

Ethics committee/IRB of Ethical Committee Research UZ/KU Leuven gave ethical approval for this work (protocol S59482), and all participants provided informed consent.

