## Supplementary Material for "Functional variation in human Carbohydrate-Active enZYmes (hCAZymes) in relation to the efficacy of a FODMAP-reducing diet in IBS patients"

### TABLES

**Table S1.** hCAZyme variants identified in this study

| Gene | Variant | BP (GRCh 38) | dbSNP | Ref allele | Alt allele | gnomAD freq | freq this study | M-CAP | M-CAP pathogenicity | CADD | CADD pathogenicity | Hypomorphic | Carriers Diet (N=196) | Carriers OB (N=54) |
|---|---|---|---|---|---|---|---|---|---|---|---|---|---|---|
| ***AMY*** | | | | | | | | | | | | | | |
| | Val190Leu | chr1:103573762 | - | G | T | 0.0000009 | 0,002 | 0.139 | 1 | - | - | 1 | - | 1 |
| | Val309Ala | chr1:103575270 | rs140209167 | T | C | 0.0025 | 0.006 | 0.170 | 1 | - | - | 1 | 2 | 1 |
| | Gly319Arg | chr1:103575299 | rs140978983 | G | A | 0.0315 | 0.044 | - | - | 27.3 | 1 | 1 | 16 | 6 |
| | Arg358His | chr1:103575512 | rs150608402 | G | A | 0.0002 | 0.004 | 0.003 | 0 | - | - | 0 | 2 | - |
| | Ile406Thr | chr1:103577605 | rs151132065 | T | C | 0.0220 | 0.018 | - | - | 26.0 | 1 | 1 | 7 | 2 |
| ***LCT*** | | | | | | | | | | | | | | |
| | Ala152T | chr2:135836716 | rs114525655 | C | T | 0.0001 | 0.002 | - | - | 16.7 | 0 | 0 | - | 1 |
| | Val219Ile | chr2:135833176 | rs3754689 | C | T | 0.1350 | 0.176 | - | - | 10.1 | 0 | 0 | 66 | 14 |
| | Ile362Val | chr2:135817964 | rs4954449 | T | C | 0.9999 | 0.998 | - | - | 13.8 | 1 | 0 | 196 | 54 |
| | Phe530Leu | chr2:135817460 | - | A | G | 0.000001 | 0.002 | 0.014 | 0 | - | - | 0 | 1 | - |
| | Gly928Cys | chr2:135809565 | - | C | A | - | 0.002 | 0.127 | 1 | - | - | 1 | 1 | - |
| | Asn1035Tyr | chr2:135809244 | - | T | A | - | 0.004 | 0.135 | 1 | - | - | 1 | 2 | - |
| | Ala1096T | chr2:135809061 | rs146467199 | C | T | 0.0009 | 0.002 | 0.006 | 0 | - | - | 0 | - | 1 |
| | Thr1329Met | chr2:135807315 | rs555708380 | G | A | 0.0000 | 0.002 | 0.010 | 0 | - | - | 0 | 1 | - |
| | Ala1483Ser | chr2:135804784 | rs139591272 | C | A | 0.0009 | 0.002 | 0.015 | 0 | - | - | 0 | 1 | - |
| | Tyr1549Cys | chr2:135803947 | rs147495948 | T | C | 0.0005 | 0.004 | 0.007 | 0 | - | - | 0 | 2 | - |
| | Asn1639Ser | chr2:135798089 | rs2322659 | T | C | 0.7803 | 0.736 | - | - | 2.6 | 0 | 0 | 179 | 50 |
| ***MGAM*** | | | | | | | | | | | | | | |
| | Ile25Val | chr7:142005603 | rs61733478 | A | G | 0.00004 | 0.002 | - | - | 12.9 | 0 | 0 | 1 | - |
| | Ile28Thr | chr7:142005613 | rs201144916 | T | C | 0.0035 | 0.004 | - | - | 13.5 | 0 | 0 | 2 | - |
| | Asp391Asn | chr7:142027685 | rs184092742 | G | A | 0.0061 | 0.008 | - | - | 13.9 | 0 | 0 | 4 | - |
| | Arg401Cys | chr7:142027715 | rs188481752 | C | T | 0.0032 | 0.002 | 0.066 | 1 | - | - | 1 | 1 | - |
| | Gln404His | chr7:142027726 | rs2272330 | G | T | 0.0091 | 0.004 | - | - | 11.0 | 0 | 0 | 1 | 1 |
| | Arg637Ser | chr7:142034793 | rs190777514 | A | C | 0.0019 | 0.002 | 0.133 | 1 | - | - | 1 | 1 | - |
| | His755Tyr | chr7:142038562 | rs113689539 | C | T | 0.0001 | 0.004 | - | - | 14.5 | 0 | 0 | 1 | 1 |
| | Leu806Ile | chr7:142040764 | rs956495934 | C | A | 0.00002 | 0.002 | 0.037 | 1 | - | - | 1 | 1 | - |
| | Leu854Phe | chr7:142047846 | rs200141280 | C | T | 0.0013 | 0.002 | 0.029 | 1 | - | - | 1 | - | 1 |

| | | | | | | | | | | | | | | |
|---|---|---|---|---|---|---|---|---|---|---|---|---|---|---|
| | Asn858Asp | chr7:142047858 | rs2960746 | A | G | 0.0001 | 0.006 | - | - | 21.2 | 1 | 1 | 3 | - |
| | Cys873Trp | chr7:142050266 | rs768578658 | T | G | 0.0001 | 0.002 | 0.059 | 1 | - | - | 1 | 1 | - |
| | Thr907Met | chr7:142050779 | rs187898444 | C | T | 0.0117 | 0.010 | - | - | 0.2 | 0 | 0 | 2 | 3 |
| | Glu976Ala | chr7:142052415 | rs116536012 | A | C | 0.0258 | 0.030 | - | - | 10.6 | 0 | 0 | 10 | 5 |
| | Arg1039His | chr7:142052941 | rs139662456 | G | A | 0.0001 | 0.002 | - | - | 7.5 | 0 | 0 | - | 1 |
| | Pro1057Ser | chr7:142054763 | rs780243925 | C | T | 0.00001 | 0.002 | 0.007 | 0 | - | - | 0 | 1 | - |
| | Pro1424Thr | chr7:142063511 | rs185053832 | C | A | 0.0108 | 0.010 | - | - | 25.0 | 1 | 1 | 3 | 2 |
| | Ile1783Val | chr7:142103290 | rs140217455 | A | G | 0.0031 | 0.006 | 0.011 | 0 | - | - | 0 | 1 | 2 |
| | Leu1794Ser | chr7:142103324 | rs201177568 | T | C | 0.0010 | 0.002 | 0.049 | 1 | - | - | 1 | 1 | - |
| ***MGAM2*** | | | | | | | | | | | | | | |
| | Ille13Thr | chr7:142116911 | - | T | C | 0.000006 | 0.002 | 0.170 | 1 | - | - | 1 | - | 1 |
| | Phe327Leu | chr7:142138562 | rs6464465 | C | G | 0.3588 | 0.380 | - | - | 17.8 | 0 | 0 | 126 | 30 |
| | Glu471Lys | chr7:142143862 | rs76786761 | G | A | 0.0598 | 0.074 | - | - | 11.9 | 0 | 0 | 27 | 8 |
| | Pro590Ser | chr7:142154151 | rs73158444 | C | T | 0.0891 | 0.082 | - | - | 18.3 | 0 | 0 | 30 | 8 |
| | Asn606Ser | chr7:142154739 | rs79591013 | A | G | 0.0122 | 0.012 | - | - | 25.9 | 1 | 1 | 4 | 2 |
| | Cys619Arg | chr7:142154777 | rs774919724 | T | C | 0.0006 | 0.002 | 0.050 | 1 | - | - | 1 | - | 1 |
| | Arg682Trp | chr7:142158057 | rs567505779 | C | T | 0.0028 | 0.002 | 0.010 | 0 | - | - | 0 | 1 | - |
| | Ser745Leu | chr7:142160147 | rs114108719 | C | T | 0.0001 | 0.002 | - | - | 11.4 | 0 | 0 | - | 1 |
| | Thr1096Met | chr7:142171376 | rs111852582 | C | T | 0.00001 | 0.002 | - | - | 18.2 | 0 | 0 | 1 | |
| | Ala1138Val | chr7:142172159 | - | C | T | - | 0.002 | 0.063 | 1 | - | - | 1 | 1 | - |
| | Leu1174Met | chr7:142172723 | rs73547325 | T | A | 0.0749 | 0.052 | - | - | 25.0 | 1 | 1 | 18 | 8 |
| | Thr1326Ala | chr7:142185128 | rs201188036 | A | G | 0.0028 | 0.006 | 0.002 | 0 | - | - | 0 | 1 | 2 |
| | Pro1425Leu | chr7:142189433 | rs563508674 | C | T | 0.00005 | 0.002 | 0.003 | 0 | - | - | 0 | 1 | - |
| | Val1430Met | chr7:142189447 | rs4726494 | G | A | 0.0087 | 0.014 | 0.013 | 0 | - | - | 0 | 7 | - |
| | Gln1546Glu | chr7:142197403 | rs7776662 | C | G | 0.00002 | 0.002 | - | - | 27.0 | 1 | 1 | - | 1 |
| | Arg1589Gln | chr7:142197533 | - | G | A | 0.000003 | 0.002 | 0.010 | 0 | - | - | 0 | - | 1 |
| | Val2018Ile | chr7:142220563 | - | G | A | - | 0.016 | 0.090 | 1 | - | - | 1 | 8 | - |
| | Ser2108Asn | chr7:142220834 | rs868461091 | G | A | 0.00005 | 0.002 | 0.019 | 0 | - | - | 0 | 1 | - |
| | Ile2384Phe | chr7:142221661 | rs114133571 | A | T | 0.0002 | 0.002 | - | - | 11.2 | 0 | 0 | 1 | - |
| | Pro2390Leu | chr7:142221680 | rs60502652 | C | T | 0.0861 | 0.080 | - | - | 11.9 | 0 | 0 | 32 | 8 |

| | | | | | | | | | | | | | | |
|---|---|---|---|---|---|---|---|---|---|---|---|---|---|---|
| ***SI*** | | | | | | | | | | | | | | |
| | Met1Ile | chr3:165076010 | - | C | T | - | 0.002 | 0.841 | 1 | - | - | 1 | 1 | - |
| | Val15Phe | chr3:165075970 | rs9290264 | C | A | 0.2990 | 0.340 | - | - | 24.5 | 1 | 1 | 113 | 30 |
| | Glu40Gly | chr3:165074667 | rs747623135 | T | C | 0.00002 | 0.002 | 0.012 | 0 | - | - | 0 | 1 | - |
| | Ser186Pro | chr3:165067419 | rs142447888 | A | G | 0.0003 | 0.002 | 0.010 | 0 | - | - | 0 | 1 | - |
| | Thr231Ala* | chr3:165065377 | rs9283633 | T | C | 0.5889 | 0.080 | - | - | 12.8 | 0 | 0 | - | - |
| | Val371Met | chr3:165059937 | rs138434001 | C | T | 0.0032 | 0.004 | 0.412 | 1 | - | - | 1 | 2 | - |
| | Arg774Gly | chr3:165038006 | rs147207752 | T | C | 0.0015 | 0.004 | 0.113 | 1 | - | - | 1 | 2 | - |
| | Ile799Val | chr3:165037931 | rs150246328 | T | C | 0.0042 | 0.004 | 0.058 | 1 | - | - | 1 | 1 | 1 |
| | Tyr975His | chr3:165023746 | rs146785675 | A | G | 0.0055 | 0.002 | - | - | 25.1 | 1 | 1 | 1 | - |
| | Ser1490Ile | chr3:164998611 | rs376437234 | C | A | 0.0001 | 0.002 | 0.053 | 1 | - | - | 1 | - | 1 |
| | Met1523Ile | chr3:164996744 | rs4855271 | C | T | 0.9072 | 0.008 | - | - | 9.6 | 0 | 0 | 196 | 53 |
| | Leu1520Ter | chr3:164996758 | - | - | A | - | 0.008 | - | - | - | - | 1 | 1 | 3 |
| ***TREH*** | | | | | | | | | | | | | | |
| | Lys140Arg | chr11:11866288 | rs34978247 | T | C | 0.0069 | 0.006 | - | - | 20.6 | 1 | 1 | 3 | - |
| | Ile328Thr | chr11:11866065 | rs200440695 | A | G | 0.0052 | 0.004 | 0.004 | 0 | - | - | 0 | 2 | - |
| | Thr389Ala | chr11:11865990 | rs2276065 | T | C | 0.2298 | 0.210 | - | - | 1.1 | 0 | 0 | 64 | 24 |
| | Tyr449His | chr11:118659457 | rs11827611 | A | G | 0.00005 | 0.004 | - | - | 4.8 | 0 | 0 | - | 2 |
| | Arg486Trp | chr11:11865899 | rs2276064 | G | A | 0.0129 | 0.008 | - | - | 16.7 | 0 | 0 | 4 | - |
| | Phe492Leu | chr11:118658976 | rs374204865 | A | G | 0.0001 | 0.002 | 0.025 | 0 | - | - | 0 | - | 1 |
| | Met512Thr | chr11:11865891 | rs556006762 | A | G | 0.0003 | 0.002 | 0.117 | 1 | - | - | 1 | 1 | - |
| | His566Tyr | chr11:11865834 | rs200772007 | G | A | 0.0005 | 0.004 | 0.004 | 0 | - | - | 0 | 1 | 1 |

**Gene:** gene name; **Variant:** corresponding amino acid change; **BP (GRCh38):** genomic position, based on human genome reference GRCh38; **dbSNP:** dbSNP database ID; **Ref Allele:** reference allele according to gnomAD v3.1.1 data (https://gnomad.broadinstitute.org/); **Alt Allele:** alternative allele according to gnomAD; **gnomAD freq:** allele frequency in individuals of non-Finnish European ancestry, as from gnomAD; **freq this study:** allele frequency in 250 IBS DOMINO samples from dietary and medication arm; **M-CAP:** Mendelian Clinically Applicable Pathogenicity (M-CAP) pathogenicity likelihood scores (http://bejerano.stanford.edu/mcap/); **M-CAP pathogenicity:** whether the variant is predicted to be pathogenic (1) or benign (0) based on M-CAP pathogenicity score >0.025; **CADD:** Combined Annotation Dependent Depletion (CADD) phred-like scores (https://cadd.gs.washington.edu/); **CADD pathogenicity:** whether the variant is predicted to be pathogenic (1) or benign (0) based on CADD pathogenicity score >20; **Hypomorphic**: whether the variant is predicted to be pathogenic (1) or benign (0) based on MCAP and CADD; **Carriers Diet**: number of carriers among 196 IBS patients from the dietary arm; **Carriers OB**: number of carriers among 54 IBS-D patients from the OB arm. **"-":** data not available. * Thr231Ala was detected but failed to generate reliable results (not adequately covered), though this is irrelevant to the aim of this study (this is not a hypomorphic variant).

| **Table S2.** Demographics and symptom scores in hCAZyme carrier groups from the dietary arm | | | |
|---|---|---|---|
| | Carriers | Non-carriers | P value |
| Patients: N | 137 | 59 | |
| Females: N (%) | 102 (74.5) | 45 (76.3) | ns |
| Age: mean ± SD | 40.8 ± 14.5 | 41.5 ± 15.2 | ns |
| **Stool type** | | | |
| IBS-D: N (%) | 34 (24.8) | 21 (35.6) | ns |
| IBS-M: N (%) | 50 (36.5) | 23 (39.0) | ns |
| IBS-C: N (%) | 31 (22.6) | 8 (13.6) | ns |
| IBS-U: N (%) | 22 (16.1) | 7 (11.9) | ns |
| **Symptoms at baseline** | | | |
| IBS-SSS score: mean ± SD | 272.3 ± 90.7 | 265.9 ± 102.1 | ns |
| Abdominal distention severity: mean ± SD | 53.0 ± 26.3 | 54.2 ± 25.3 | ns |
| Life disruption: mean ± SD | 49.8 ± 25.6 | 52.5 ± 27.4 | ns |
| Abdominal pain severity: mean ± SD | 50.4 ± 25.2 | 47.7 ± 23.6 | ns |
| Bowel habit satisfaction: mean ± SD | 68.7 ± 23.0 | 64.6 ± 24.3 | ns |
| Abdominal pain duration (x10 days): mean ± SD | 4.9 ± 2.9 | 4.9 ± 3.1 | ns |

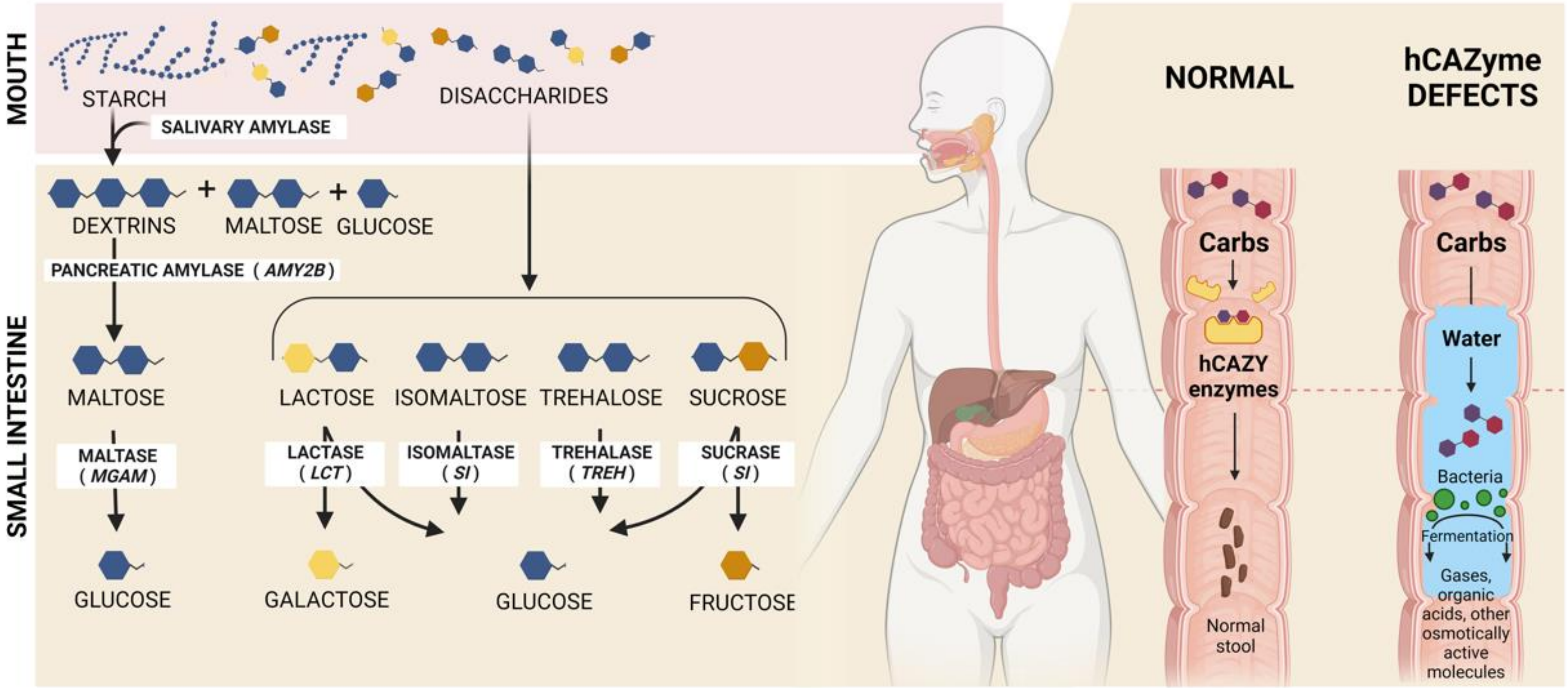

**Figure S1.** Graphical representation of the process of carbohydrate digestion in the gastrointestinal system, including defects in hCAZYmes.

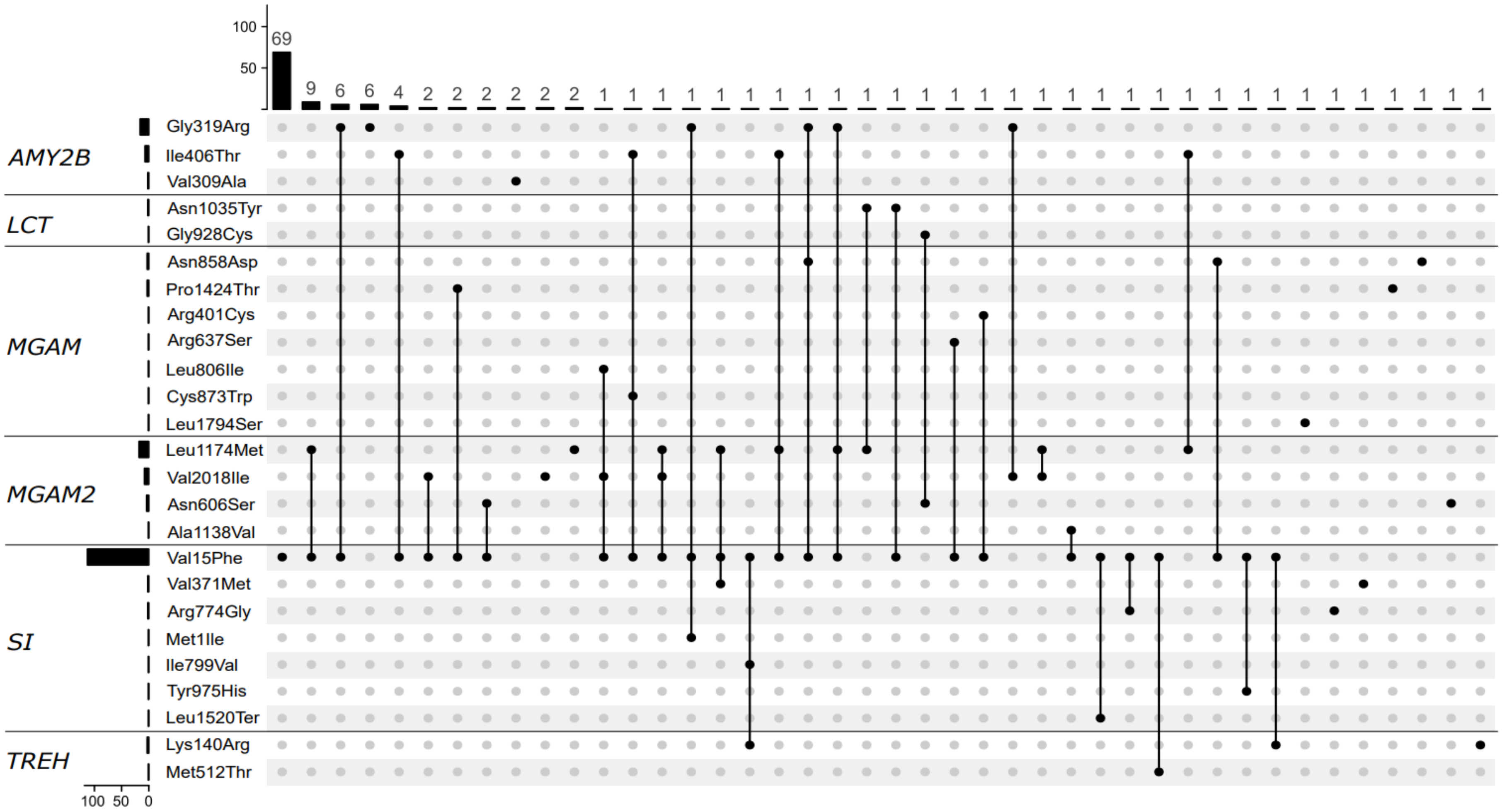

**Figure S2.** UpSet plot showing all genotypic combinations of hypomorphic variants present in hCAZyme genes from this study.

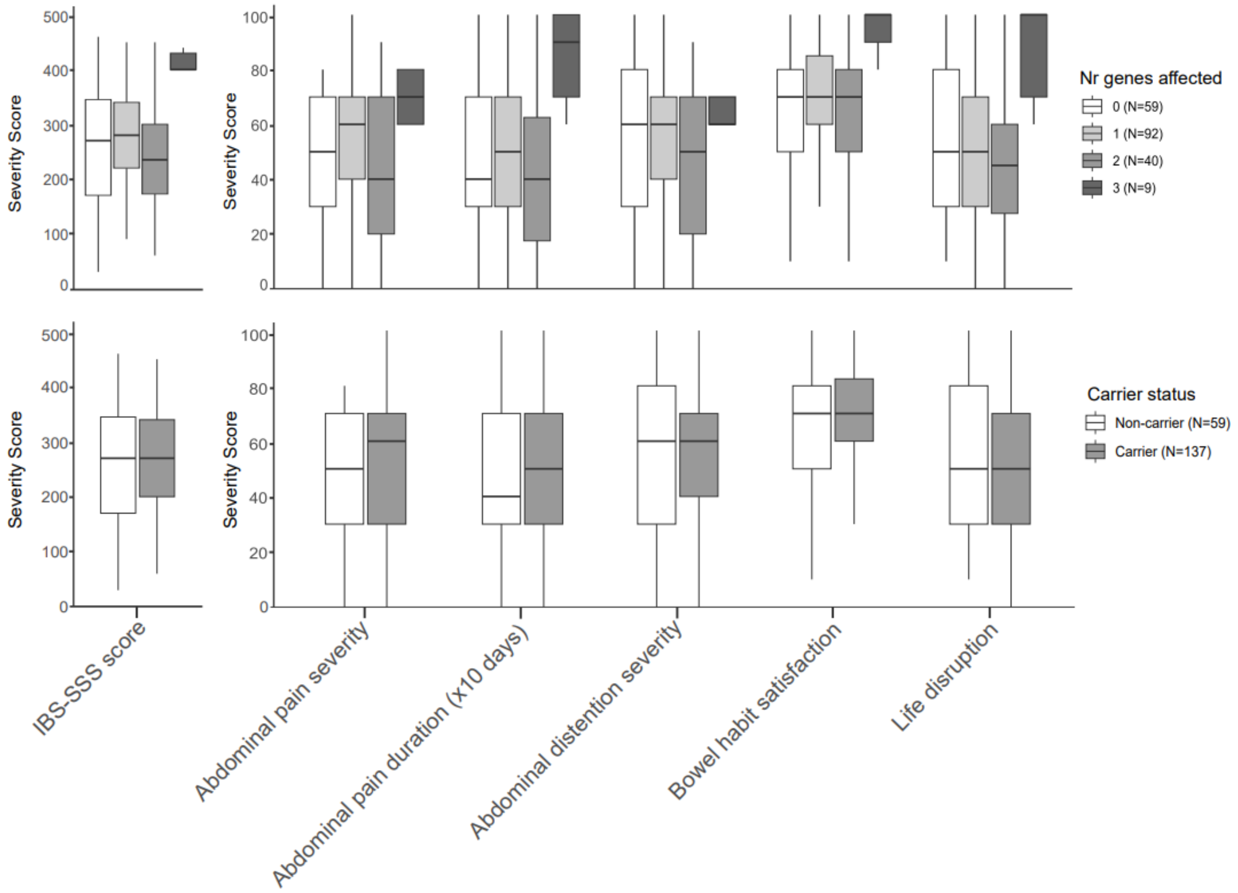

**Figure S3.** Baseline IBS symptoms in 196 IBS DOMINO patients from the dietary arm stratified according to genetic variation in the hCAZyme genes: top) number of affected hCAZyme genes; bottom) hCAZyme carrier status.

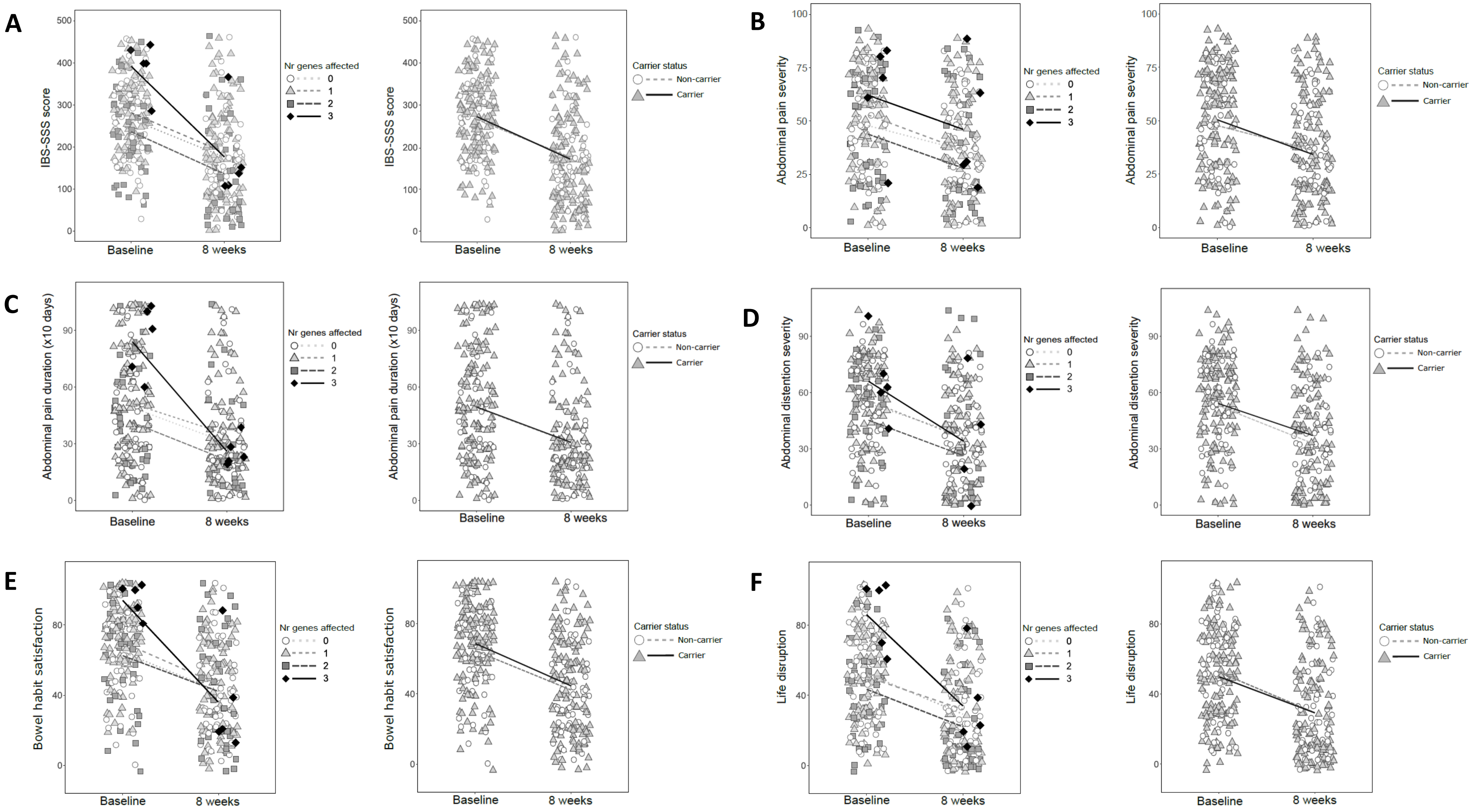

**Figure S4.** Variation of IBS symptoms after 8 weeks of treatment in 196 IBS patients from the dietary arm. Total (A) and individual (B-F) symptom scores are stratified according to genetic variation in the hCAZyme genes: left) number of affected hCAZyme genes; right) hCAZyme carrier status.

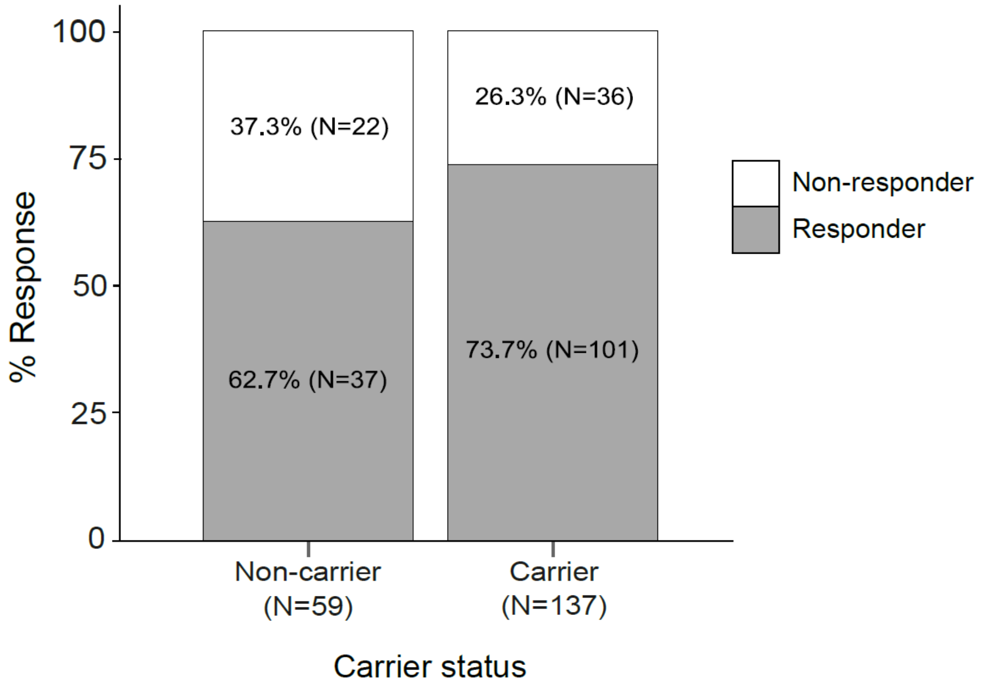

**Figure S5.** Response to a FODAMP-lowering diet in 196 IBS DOMINO patients stratified according to hCAZyme carrier status.

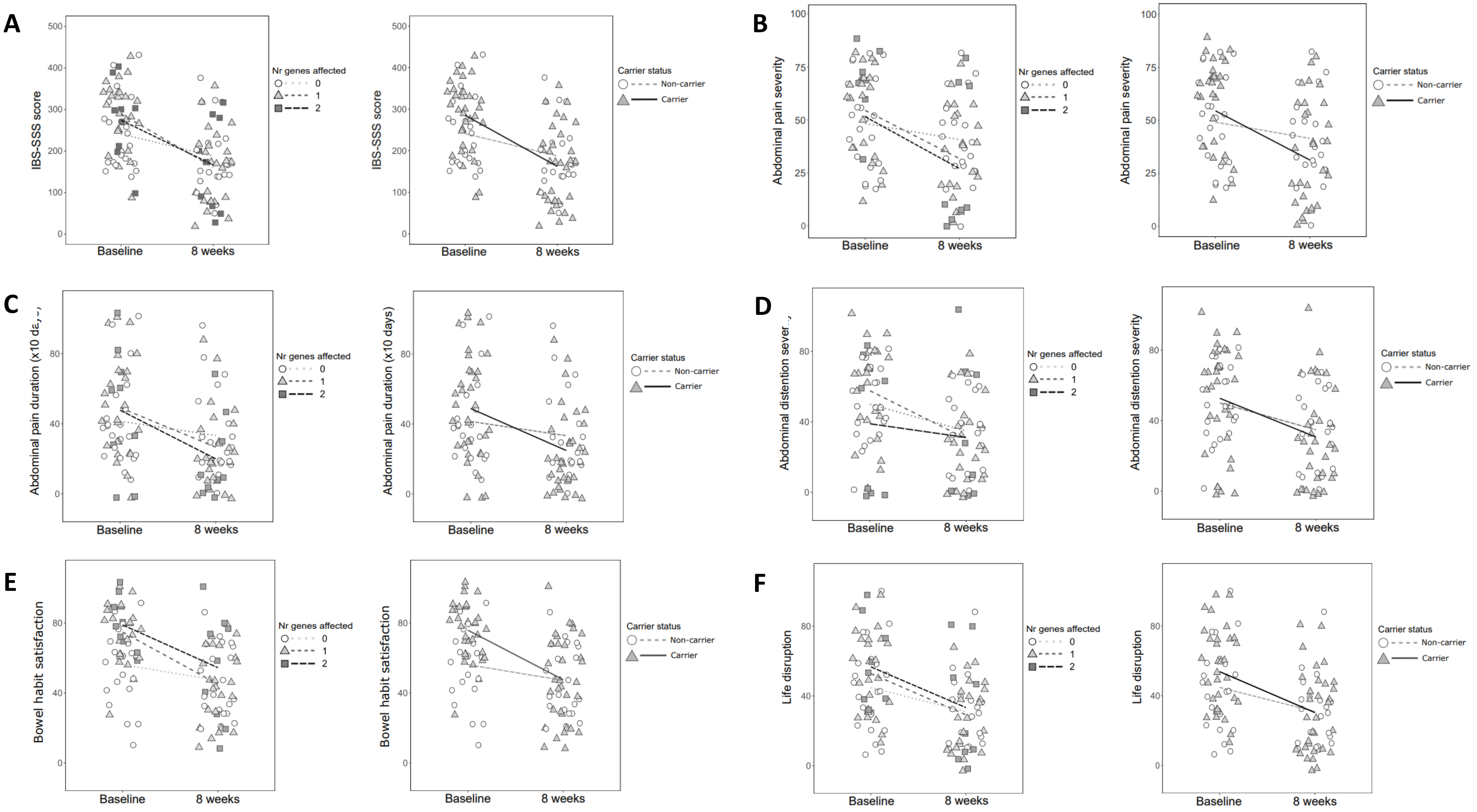

**Figure S6.** Variation of IBS symptoms after 8 weeks of treatment in 55 IBS-D patients from the dietary arm. Total (A) and individual (B-F) symptom scores are stratified according to genetic variation in the hCAZyme genes: left) number of affected hCAZyme genes; right) hCAZyme carrier status.

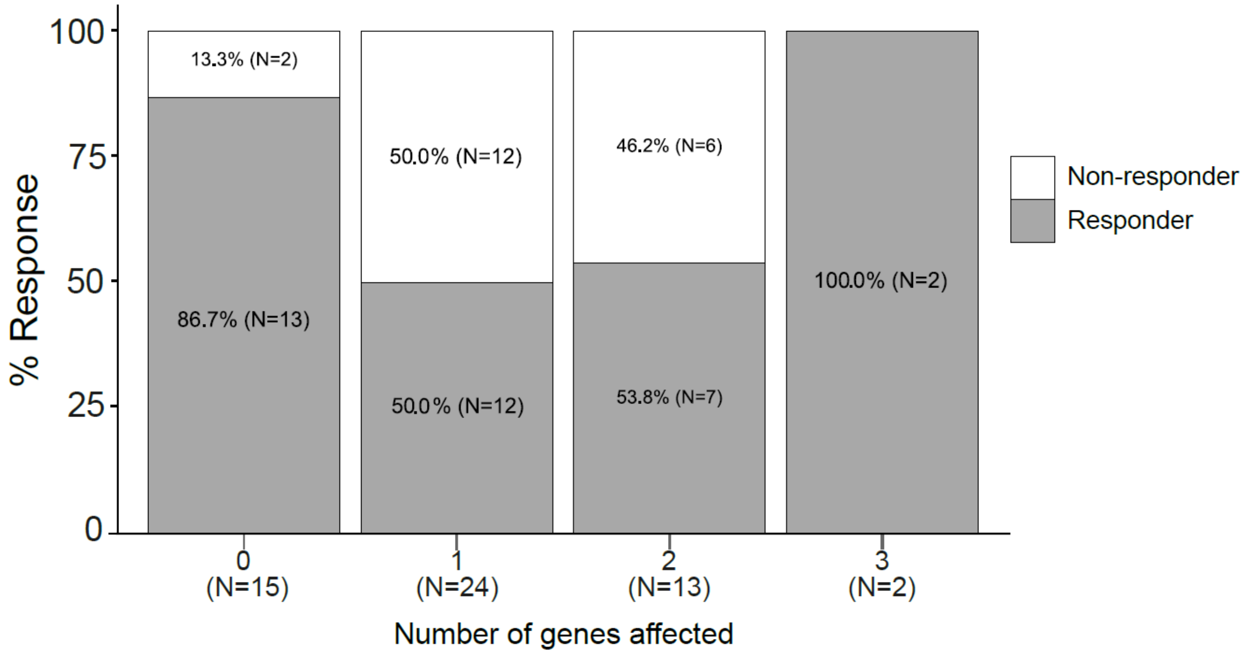

**Figure S7.** Response to OB treatment in 54 IBS-D IBS DOMINO patients stratified according to the number of hCAZyme genes affected.
